# Social Norms and Security and Justice Services for Gender-based Violence in Nepal: Programmatic Implications from a Baseline Mixed-Methods Assessment

**DOI:** 10.1101/2024.01.20.24300968

**Authors:** Cari Jo Clark, Brian Batayeh, Iris Shao, Irina Bergenfeld, Manoj Pandey, Sudhindra Sharma, Shikha Shrestha, Amritha Gourisankar, Anudeeta Gautam, Tehnyat J. Sohail, Holly Shakya, Grace Morrow, Abbie Shervinskie, Subada Soti

## Abstract

**Background:** Gender-based violence (GBV) is highly prevalent throughout the world. Only a small fraction of survivors seek help from security and justice providers (S&J) such as the police or courts, due in part to social norms that discourage help-seeking. Social norms interventions have proven effective in combatting GBV but have not been tested alongside S&J interventions. The Strengthening Access to Holistic, Gender Responsive, and Accountable Justice in Nepal (SAHAJ) project was designed to fill this gap. We provide a comprehensive mixed-methods situation analysis of GBV-related social norms, help-seeking, S&J service provision.

**Methods:** The SAHAJ evaluation is a 3-armed, mixed-methods evaluation of the impact of social norms programming alone, S&J programming alone, and combined programming in 17 communities in Nepal. Baseline data included a household survey (N=3830), a sub-study of youth (N=143) and married adults (N=464) in one site and qualitative data collection including interviews with S&J service providers, help-seeking GBV survivors and families (N=68), and focus group discussions with police, youth groups, and school management committees (N=20) in four sites. Descriptive analysis of survey data was triangulated with findings from a modified grounded theory analysis of the qualitative data to elucidate the role of social norms and other barriers limiting help-seeking.

**Results:** GBV was perceived to be common, especially child marriage, domestic violence, eve-teasing, and dowry-related violence. Formal help-seeking was low, despite positive attitudes towards S&J providers. Participants described injunctive norms discouraging formal reporting in cases of GBV and sanctions for women violating these norms.

**Conclusions:** Norms favoring family- and community-based mediation remain strong. Sanctions for formal reporting remain a deterrent to help-seeking. Leveraging gender-equitable role models, such as female S&J providers, and connecting S&J providers to women and youth may capitalize on existing shifts.

## Introduction

Gender-based violence is any harmful act directed at an individual or group based on their gender. GBV includes, but is not limited to, intimated partner violence (IPV), in-law abuse, sexual assault, dowry-related violence, child marriage, and sexual harassment or assault, with IPV being the most pervasive. Globally, approximately 1 in 3 women have experienced physical or sexual intimate partner violence (IPV),(1) but less than 9% of survivors have sought help from the police,(2) a prominent actor in the delivery of services meant to safeguard personal safety and security and uphold the rule of law. This is despite decades of legal and procedural improvements: currently 85% of the 190 countries surveyed by the World Bank’s Women, Business, and the Law project, have criminalized domestic violence.(3) Over this time, much has been accomplished, including a shift toward survivor-centered, gender sensitive services, improved legal proceedings, and outcome enforcement. However, these improvements are modest relative to the investment, in part due to social norms that discourage disclosure outside the family and the interference of others in private affairs.

Interventions designed to change norms can positively influence individual attitudes and practices around gender-based violence (GBV). Multicomponent interventions have shown increased knowledge and awareness about IPV,(4, 5) decreased endorsement of gender-inequitable attitudes,(4–7) increased joint household decision-making,(5) and the reduction of violence against women,(7–9) including female genital cutting.(10) There is some evidence that norms-focused interventions can reduce harmful norms,(4, 11) improve confidence in service providers,(11) and help-seeking,(4) although none have examined the potential impact of programming on help-seeking from security and justice (S&J) providers.

To begin to fill this gap in S&J and social norms-focused interventions, the Strengthening Access to Holistic, Gender Responsive, and Accountable Justice in Nepal (SAHAJ) project was launched as an integrated multi-component S&J and social norms-focused intervention targeting families, schools, and S&J providers as part of the United Kingdom’s Government’s Integrated Programme for Strengthening Security and Justice (IP-SSJ). **We provide a comprehensive mixed-methods situation analysis of GBV-related social norms, help-seeking, S&J service provision.**

## Background

### GBV in Nepal

In Nepal, an estimated 23% of reproductive age women report having experienced physical violence victimization since age 15 and 8% report sexual violence victimization in their lifetime, with the Madhesh Province having the highest prevalence estimates.(12) If traditional practices such as *chhaupadi* (isolation of women and girls during menstruation and just after childbirth) and child marriage are considered, the prevalence estimates would exceed 80% in areas where these practices are endemic. Despite progress over the past few decades, such as making IPV, *chhaupadi*, and child marriage punishable offenses, as well as greater inclusion of women in the political realm and increased education for girls,(13, 14) women in Nepal often experience limitations in their ability to seek education, employment, or socialize freely outside the home. Nepali men tend to be dominant in household decision-making, ascribe to traditional gender roles, and hold significant control over their wives. The acceptability of IPV and other form of GBV remains relatively high among Nepali men and women, alike.(12, 15, 16)

### S&J sector in Nepal

The S&J sector in Nepal is a mixture of formal and informal services. Indigenous practices of mediation or adjudication by local elders and leaders, the oldest of these informal services, are easily accessible, efficient, and a means to preserve family honor and social harmony.(17, 18) However, these practices are male-dominated and prioritize social harmony over individual needs, i.e., they are not survivor-centered and may be politicized, especially when justice is facilitated by persons affiliated with a political party. Formal S&J actors (police, court system, lawyers) are a more recent development, but there have been setbacks in establishing their utility, including accusations of political interference; corruption; discrimination; physical, financial, and social inaccessibility; and a lack of timely resolution, especially for the judiciary.(17–23) Despite these challenges, considerable reforms in infrastructure, equipment, and professionalism are underway. Importantly, these reforms have begun among the police who have become a preferred source of assistance for crime, especially for domestic violence cases in the Madhesh and Lumbini Provinces (formerly Province 2 and Province 5, respectively) where the present study is set.(24, 25) A special Directorate of the police was established to address a lack of professional skills within the police force to deal with GBV and related crimes against women, children, and elders, including gender responsive and victim centered approaches.(26) While well-regarded, its reach into the community remains less than that of the traditional police force. After the 2017 elections, Judicial Committees were established to bring judicial services closer to the community as part of the decentralization process. These committees are headed by the deputy mayor of urban municipalities and the vice-chairperson of rural municipalities, newly formed posts through the government restructuring process headed predominantly by women—an important opportunity to support female leadership development. The committees, while established throughout the country, have been criticized for insufficient legal training to effectively address GBV and are not yet fully resourced to perform their mandate.(27) This legal capacity deficit is not unique to the Judicial Committees, but a broader challenge arising from the devolution of governance to the local level.(28)

### S&J Help-seeking for GBV

In Nepal, as in many settings around the world, a general lack of help-seeking for GBV predominates.(29) According to nationally representative data of reproductive age women, only 8.7% of women who had experienced physical or sexual violence sought help from formal sources in 2022, with the police being the most common (7.2%). Instead, these women more often turned to informal sources, such as their maternal family (63%) and neighbors (35%), to help end the violence.(15) Non-disclosure has been linked to a desire to maintain family honor and privacy, fear of disrupting family relations and further abuse, and limited faith in the justice system.(30, 31) Similarly, poor and marginalized communities do not seek police and justice support as they perceive services as discriminatory based on ethnicity, caste, income level, and gender.(18) A woman’s decision to seek justice is heavily dependent on the views of her family and community, the severity of her problems, and an often-inadequate knowledge of available services.(32) Many view the police as a last resort or to be used only for serious cases and many cases of GBV are not considered serious enough to warrant police intervention.(33)

## Methodology

### Overview

This study is a concurrent mixed-methods baseline assessment of 17 toles distributed across 3 study arms in 9 districts located in the Madhesh and Lumbini provinces. One arm contained only family- and school-based social norms approaches (N=4 sites). A second arm included only S&J activities (N=8 sites) designed to improve gender responsiveness, victim-centeredness, and social accountability among S&J service providers. The third arm contained a combination of S&J family and school-based social norms approaches (N=5 sites). Data presented here are from the baseline assessment (14-08-2019 and 07-09-2019). The study was approved by the Nepal Health Research Council (#602/2019) and the Institutional Review Board at Emory University (IRB00110703). All participants provided written informed consent, signing with an X if non-literate and co-signed by the interviewer.

### Site selection

The project was located in municipalities which had undergone or were undergoing S&J-focused infrastructure improvement and initiation of community policing. Project activities were administered within wards (geographic units smaller than municipalities) based on the proportion of marginalized and vulnerable groups from a systematic fieldwork process undertaken by project implementers. Social norms activities were located within toles, which are sub-divisions of wards. Tole selection involved discussions with local government, police, non-governmental organizations, and committees tasked with GBV prevention and response to identify communities in particular need of programming. Additional considerations were the presence of secondary schools for the school-based activities and the absence of social norms programming administered by other organizations. For consistency in the S&J only arm, wards were segmented into toles and one tole randomly selected for measurement. In addition to these sites, the research team selected one tole in ward 1 of Kapilvastu municipality to serve as the site of the sub-study because of high rates of child marriage, polygamy, the practice of dowry, domestic violence, and mobility restrictions for women as reported by project implementing partners. For the qualitative data collection, 2 sites in each province were selected to maximize variation(34) within the combined approach arm.

### Sampling and eligibility

We conducted a household survey among the toles selected for measurement across arms. The household listing included approximately 225 households in each locality. Sites that were larger than this size were segmented and those that were smaller were combined with adjacent toles to obtain the required sample size. Within each household, a married woman 18 years or older, or if not available, any knowledgeable adult 18 years or older (N=3830; 99.7% female) was administered the household survey. For the sub-study within the Lumbini Province, we additionally surveyed all adolescents aged 15 to 19 or in grades 9 to 12 (male=75; female=68) and married adults (men N=221; women N=243). Sociodemographic characteristics of the respondents in these surveys is presented in **Table 1**. Key differences across the respondent groups include the predominance of women respondents in the household survey compared to a more equal representation of male and female respondents in the sub-study and age differences with the youth survey participants being the youngest. These differences reflect the sampling strategy. The youth sub-study respondents are also more educated on average than the adult respondents, which reflects a well-established trend in Nepal and the predominance of religious minorities in the sub-study reflects the socio-demographic characteristic of the district which proportionally contains more Muslims than other districts.

**Table 1:**
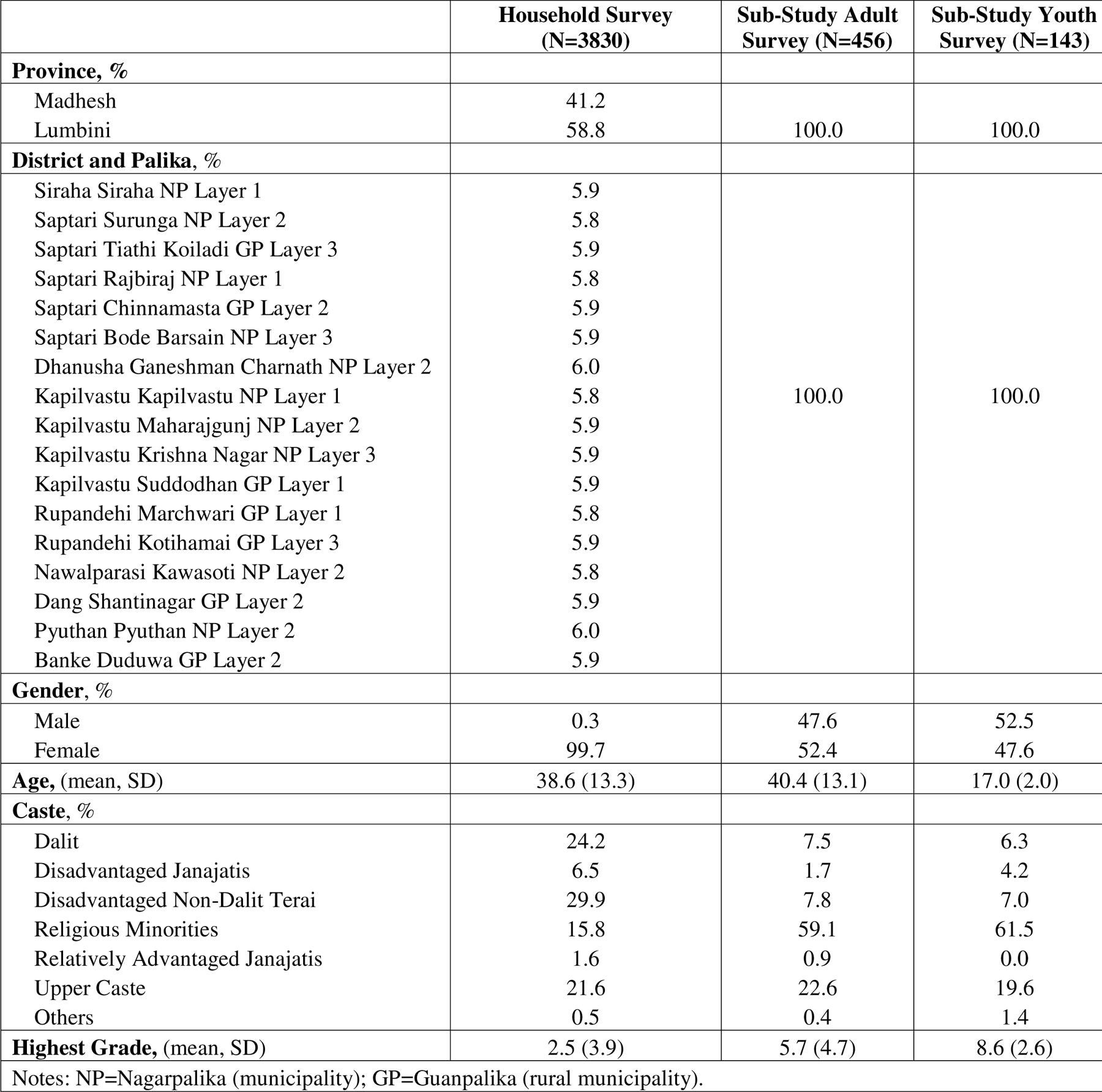
Sample size and socio-demographics, by survey type.

|  | **Household Survey**<br>**(N=3830)** | **Sub-Study Adult**<br>**Survey (N=456)** | **Sub-Study Youth**<br>**Survey (N=143)** |
| --- | --- | --- | --- |
| **Province, %** |  |  |  |
| Madhesh | 41.2 |  |  |
| Lumbini | 58.8 | 100.0 | 100.0 |
| **District and Palika, %** |  |  |  |
| Siraha Siraha NP Layer 1 | 5.9 |  |  |
| Saptari Surunga NP Layer 2 | 5.8 |  |  |
| Saptari Tiathi Koiladi GP Layer 3 | 5.9 |  |  |
| Saptari Rajbiraj NP Layer 1 | 5.8 |  |  |
| Saptari Chinnamasta GP Layer 2 | 5.9 |  |  |
| Saptari Bode Barsain NP Layer 3 | 5.9 |  |  |
| Dhanusha Ganeshman Charnath NP Layer 2 | 6.0 |  |  |
| Kapilvastu Kapilvastu NP Layer 1 | 5.8 | 100.0 | 100.0 |
| Kapilvastu Maharajgunj NP Layer 2 | 5.9 |  |  |
| Kapilvastu Krishna Nagar NP Layer 3 | 5.9 |  |  |
| Kapilvastu Suddodhan GP Layer 1 | 5.9 |  |  |
| Rupandehi Marchwari GP Layer 1 | 5.8 |  |  |
| Rupandehi Kotihamai GP Layer 3 | 5.9 |  |  |
| Nawalparasi Kawasoti NP Layer 2 | 5.8 |  |  |
| Dang Shantinagar GP Layer 2 | 5.9 |  |  |
| Pyuthan Pyuthan NP Layer 2 | 6.0 |  |  |
| Banke Duduwa GP Layer 2 | 5.9 |  |  |
| **Gender, %** |  |  |  |
| Male | 0.3 | 47.6 | 52.5 |
| Female | 99.7 | 52.4 | 47.6 |
| **Age, (mean, SD)** | 38.6 (13.3) | 40.4 (13.1) | 17.0 (2.0) |
| **Caste, %** |  |  |  |
| Dalit | 24.2 | 7.5 | 6.3 |
| Disadvantaged Janajatis | 6.5 | 1.7 | 4.2 |
| Disadvantaged Non-Dalit Terai | 29.9 | 7.8 | 7.0 |
| Religious Minorities | 15.8 | 59.1 | 61.5 |
| Relatively Advantaged Janajatis | 1.6 | 0.9 | 0.0 |
| Upper Caste | 21.6 | 22.6 | 19.6 |
| Others | 0.5 | 0.4 | 1.4 |
| **Highest Grade, (mean, SD)** | 2.5 (3.9) | 5.7 (4.7) | 8.6 (2.6) |
| Notes: NP=Nagarpalika (municipality); GP=Guanpalika (rural municipality). |  |  |  |

In the four sites identified for qualitative data collection (**Table 2**), we held focus group discussions (FGDs) with adolescents in youth clubs (N=8 groups, 70 individuals), ward police (N=4 groups, 33 individuals), members of the school management committee (N=4 groups, 28 individuals) and the women’s police cell (N=4 groups, 24 individuals).We also conducted in-depth interviews (IDIs) with formal and informal S&J providers (police chief, women’s police cell, GBV control group member, and local government officials including a judicial committee representative) (N=19), GBV help-seeking survivors (N=4), school administrators (N=4). and 12 families (N=41 individuals) recruited into SAHAJ family-based programming: The husband, wife, mother-in-law, and father-in-law of each family were interviewed in the Madhesh Province, and the mother, father, and adolescent daughter of each family in the Lumbini Province. This slight difference in family members reflected the focus of each province, which was married women’s safety and help-seeking in the Madhesh Province and adolescent girls’ safety and help-seeking in the Lumbini Province. **Table 2** summarizes the qualitative data collected within each site.

**Table 2:** Qualitative sample, by data collection site.

| Province | District | Palika | Ward | Qualitative Sample |
| --- | --- | --- | --- | --- |
| Madhesh | Siraha | Siraha NP | 1 | FGDs: 1) youth group (female, N=1 group, 7 individuals); 2) youth group (male, N=1 group, 7 individuals); 3) School Management Committee (N=1 group, 9 individuals); 4) ward police (N=1 group, 8 individuals); 5) Women's Cell (N=1 group, 6 individuals). IDIs: 1) police chief (N=1); 2) Representative of the Women's Cell (N=1); 2) Gender-based Violence Control Group member (N=1); 3) Judicial Committee member (N=1); 4) Deputy mayor (N=1); 5) Help-seeking Survivor (N=1); 7) School administrator (N=1); 8) 3 families consisting of a husband, wife, wife's mother-in-law, wife's father-in-law (N=12). |
|  | Saptari | Rajbiraj NP | 6 | FGDs: 1) youth group (female, N=1 group, 8 individuals); 2) youth group (male, N=1 group, 8 individuals); 3) School Management Committee (N=1 group, 9 individuals); 4) ward police (N=1 group, 11 individuals); 5) Women's Cell (N=1 group, 6 individuals). IDIs: 1) police chief (N=1); 2) Representative of the Women's Cell (N=1); 3) Gender-based Violence Control Group member (N=1); 4) Judicial Committee member (N=1); 5) Deputy mayor (N=1); 6) Help-seeking Survivor (N=1); 7) School administrator (N=1); 8) 3 families consisting of a husband, wife, wife's mother-in-law, wife's father-in-law (N=12). |
| Lumbini | Kapilvastu | Kapilvastu NP | 1 | FGDs: 1) youth group (female, N=1 group, 10 individuals); 2) youth group (male, N=1 group, 10 individuals); 3) School Management Committee (N=1 group, 5 individuals); 4) ward police (N=1 group, 7 individuals); 5) Women's Cell (N=1 group, 6 individuals). IDIs: 1) police chief (N=1); 2) Representative of the Women's Cell (N=1); 3) Gender-based Violence Control Group member (N=1); 4) Judicial Committee member (N=1); 5) Help-seeking Survivor (N=1); 6) School administrator (N=1); 7) 3 families consisting of a father, mother and adolescent girl, except for 1 family where the father was unavailable for interview (N=8). |
|  | Rupandehi | Marchwari GP | 3 | FGDs: 1) youth group (female, N=1 group, 11 individuals); 2) youth group (male, N=1 group, 9 individuals); 3) School Management Committee (N=1 group, 5 individuals); 4) ward police (N=1 group, 7 individuals); 5) Women's Cell (N=1 group, 6 individuals). IDIs: 1) police chief (N=1); 2) Representative of the Women's Cell (N=1); 3) Gender-based Violence Control Group member (N=1); 4) Judicial Committee member (N=1); 5) Deputy mayor (N=1); 6) Help-seeking Survivor (N=1); 7) School administrator (N=1); 8) 3 families consisting of a father, mother and adolescent girl (N=9). |

### Data

Data collected was complementary across qualitative and quantitative data collection tools (**Table 3**). The qualitative and quantitative tools can be found in the **Online Supplement**.

**Table 3.** Measurement topics by data collection tool.

| ***Instrument*** | ***Purpose*** |
| --- | --- |
| Household survey | To collect baseline measures of sociodemographics, attitudes, descriptive and injunctive norms, and perceptions of S&J actors. |
| In-depth interviews with S&J providers and help-seeking survivors | To assess the service context and experience seeking help. |
| In-depth interviews with families | To assess decision-making, descriptive and injunctive norms, violence experience, help-seeking, and engagement with committees involved in GBV prevention or response. |
| Focus group discussions | To identify norms, norms reference groups, sanctions, sensitivity to sanctions with particular reference to family privacy, acceptability of GBV, acceptability of help seeking for GBV |
| Sub-study survey | To measure attitudes, descriptive and injunctive norms, perceptions of S&J actors, exposure to violence and help-seeking. |

Descriptive norms (perceptions of prevailing practices in the community) were measured in the household survey and the sub-study survey with items assessing whether each of 7 forms of GBV were widespread in their community. For the household survey, the respondent was asked whether the statement was true or false. In the sub-study, the participant was asked to rate their level of agreement on a 4-point Likert scale from strongly agree to strongly disagree. The items were dichotomized to obtain a percent agreeing or strongly agreeing versus disagreeing or strongly disagreeing that the practice was prevalent in their community. Injunctive norms (perceptions about what are acceptable practices in the community) were measured in the household survey and the sub-study with 15 items based on the Partner Violence Norms scale(35) and an ongoing IP-SSJ evaluation of the consortium.(36) The respondent was asked to report the extent to which members of their community espoused the sentiment on a 5-point Likert scale from nearly all members of the community to no members of the community. Items were dichotomized representing perceptions that most or nearly all persons in a community held that belief.

Physical and / or sexual Intimate partner violence was measured among adult women in the sub-study with 8 items from the Demographic and Health Survey, which were also measured in the IP-SSJ evaluation. Items measured the frequency (never, sometimes, or often) of the act in the prior 12 months. A dichotomous measure was created representing exposure to any act of IPV. In-law abuse was measured among adult women in the sub-study with 2 items from a scale developed in prior research in Nepal(37) assessing exposure (yes/no) to physical violence perpetrated by the husband’s family and in-laws incitement of partner violence. A dichotomous measure was created representing exposure to either act. Child maltreatment was measured among youth in the sub-study with the 7-item Short Child Maltreatment Questionnaire developed by the World Health Organization Regional Office for Europe.(38) Each item assessed whether the act occurred never, once or twice, or many times in the past 12 months. A dichotomous measure was created representing exposure to any act. To assess help-seeking, persons reporting violence were asked whether they had told anyone about it in the past 12 months. Item response options included 17 types of informal and formal help-seeking choices and no one.

Perceptions about S&J actors were measured in the household survey and sub-study with 2 items from the IP-SSJ monitoring tool which was being used across projects in the consortium to assess whether the police are trustworthy people and whether citizens have a role to play in supporting the police to maintain law and order. Each item was assessed on a 4-point Likert scale from strongly agree to strongly disagree, including a don’t know response option. Items were dichotomized to obtain a percent agreeing or strongly agreeing versus disagreeing or strongly disagreeing. In the sub-study, participants were asked to respond to 6 questions developed for the study about each of 5 S&J actors (police, Judicial Committee, ward Chair, local leader, and women’s committee) to assess their perceptions of aspects of victim-centered services in response to a report of physical or sexual violence to that actor. Respondents were asked whether they would be willing to report the case to that actor, whether they believed the case would be taken seriously, whether the survivor would be treated with respect, whether her privacy would be protected, whether the perpetrator would be punished, and whether the survivor would experience negative social repercussions for reporting. Each question was assessed on a 4-point Likert scale from strongly agree to strongly disagree with a response option for don’t know. Items were categorized into 1) agree or strongly agree, 2) disagree or strongly disagree, 3) and don’t know.

IDI guides for S&J providers and help-seeking survivors were developed to examine the S&J service context and help-seeking with questions modeled after constructs derived from SDDirect’s Security Sector Module.(39) IDIs with families examined decision-making, descriptive and injunctive norms on violence against women and girls and help-seeking for GBV, experience of violence and help-seeking, and engagement in local violence-related committees, with questions developed for the study and vignettes modeled after CARE’s Social Norms Analysis Plot (SNAP) framework. We designed focus group discussions (FGDs) to assess norms, norms’ reference groups, sanctions, sensitivity to sanctions with particular reference to family privacy, and understand the acceptability of GBV and help seeking for GBV, with questions developed for the study including vignettes also modeled after the SNAP framework.(40)

### Analysis

Descriptive analyses (frequencies and means, minimums and maximums) of the household survey data were performed by site and province. For the sub-study, descriptive analyses were conducted by respondent type (adult man, adult woman, male adolescent and female adolescent). For qualitative data, we developed a codebook based on an initial reading of 15 transcripts and added codes over the initial stage of the analysis to incorporate emergent themes. The revised codebook was then used in 2 rounds of inter-coder reliability testing (evaluated using Cohen’s kappa(41)) among 2 team members using a subset of the transcripts. Following each round of testing, team debriefs were used to resolve discrepancies and make minor edits to codes and definitions. The team then coded the remaining transcripts. All coding, cross-classification, and inter-coder reliability testing was performed in MAXQDA 18 (Berlin, Germany). A narrative analysis of the family interviews began with a matrix summarizing key social norms and S&J themes in each interview, which was used to create summaries of each family. Thick descriptions using the entire data set were also generated with an analytic focus on key themes, interconnectedness, and synergies.

## Results

### GBV is perceived to be widespread

Descriptive norms around the extent of GBV suggest that it is perceived to be widespread with some regional and locality differences. Forms of violence most commonly reported by participants across the sample included child marriage, eve teasing, domestic violence, and dowry-related violence. Overall, a greater proportion of individuals in the Madhesh Province reported various forms of GBV to be widespread compared to the Lumbini Province, except for eve teasing (i.e. public sexual harassment or assault) which was reported to be widespread in both provinces and Chaupadhi which was reported to be widespread by 16% of the sample in the Madhesh Province and 20% of the sample in the Lumbini Province (**Figure 1**)Error! Reference source not found.. The tole minimums and maximums suggest considerable diversity within province in these perceptions (**Online Supplement Table 1)**.

**Figure 1:**
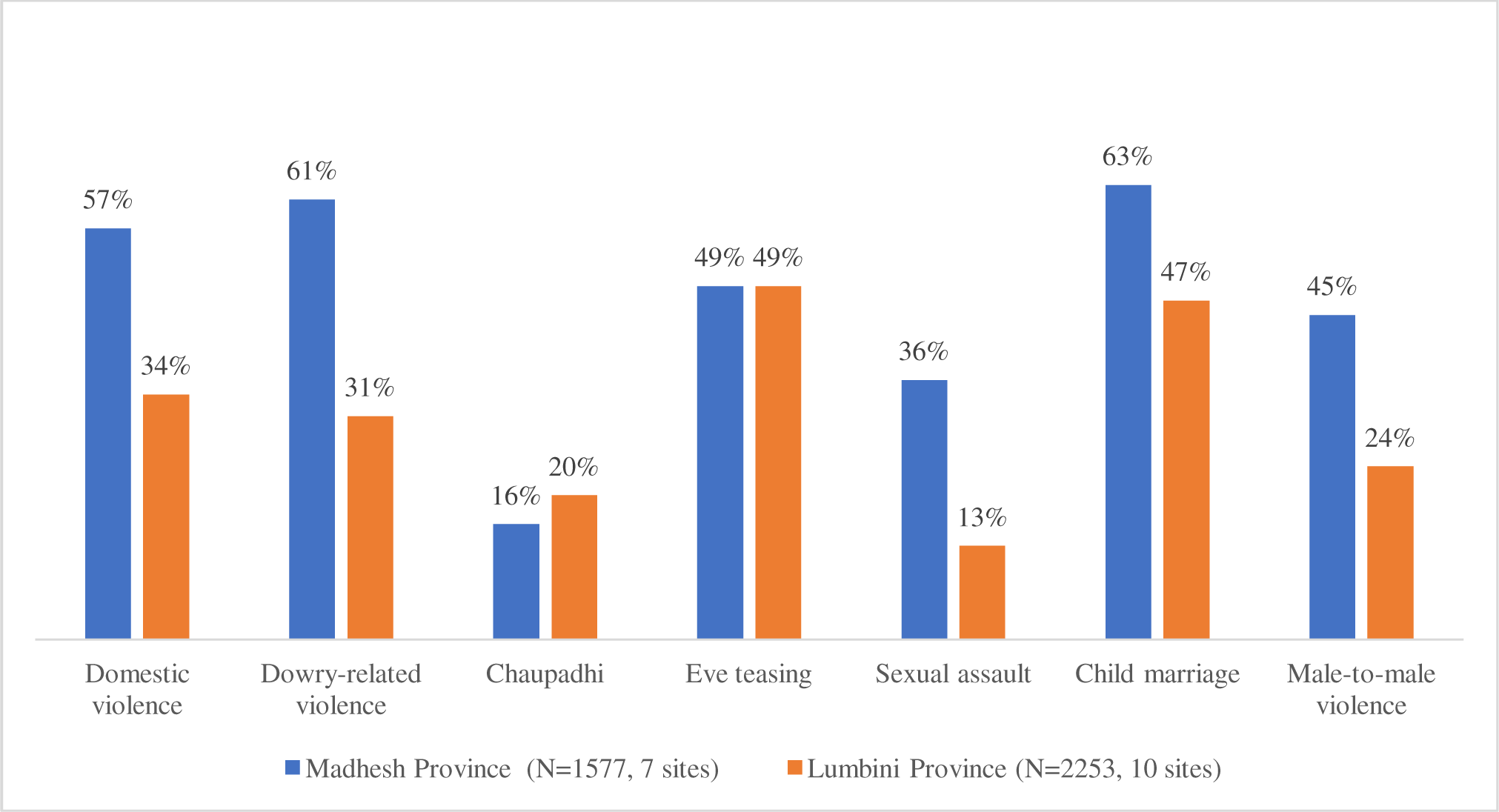
Mean percent agreement with descriptive norms items, household survey, by province.

Reports from the qualitative data also revealed that respondents characterized the acts mentioned above, as well as other acts, as GBV. IPV and abuse from in-laws were described as common across all 4 sites and took the form of beatings, verbal abuse, financial abuse (e.g. withholding of basic needs), severe restrictions on mobility, and refusal to provide necessary documents (e.g. birth and marriage certificate). Dowry issues were discussed both as a cause of GBV and a form of GBV and were implicated in both IPV and abuse from in-laws across all sites.

> *“When a daughter is married to someone after paying a dowry of 10 lakh and a motorbike, but still after reaching her in-laws house, she will be tortured for not bringing more dowries. Everyone in the family will torture her. They will rebuke her for not bringing more dowries and she will be abused by everyone… Dowry is the major reason for abusing someone.” (Saptari, S&J provider)*

Child marriage also was perceived as prevalent. Respondents partially attributed early marriage to low levels of education and awareness, financial strain on the household, reduced dowry for young brides, and concerns about the family’s reputation. According to male youth, girls who are perceived to have relationships with boys or who engage in other “bad” behavior are at risk of early marriage. Child marriage was often connected to broader discrimination against women and girls, which is common across sites and often takes the form of denying girls the same educational opportunities as boys. Participants of all types stated that girls are withdrawn from school earlier, sent to lower quality schools, burdened by household chores, face menstrual restrictions, and are provided with less nutrition while boys study or play.

### Prevalence of GBV differs by type, but common across types is a lack of help-seeking

The most frequent form of violence reported was child marriage. Based on the household survey data, 27.3% of married women were married before age 15 and 60.2% married before age 18. Help-seeking for child marriage was not assessed as the timing of the experience was not assessed. In the sub-study survey, among the adult female respondents, 4.5% (n=11) reported physical and or sexual IPV in the past 12 months. Approximately 7% (n=16) of the adult female respondents reported being hit, kicked, punched or otherwise physically hurt by a member of their husbands’ families in the past 12 months and 5.4% (n=13) reported that their in-laws encouraged their husband to do the same. Of the 9.5% (n=23) of women who reported IPV or in-law abuse, 91.3% (n=21) told no one. The remaining 2 told family or friends. Among the youth, 14.7% (N=11) of male respondents and 16.2% (N=11) of the female respondents reported child maltreatment from a parent or caretaker in the prior 12 months. Only one of the youth reported their experience of child maltreatment and they did so to family or friends.

### S&J providers, particularly police, are an acceptable source of help

**Table 4 reports perceptions of respondents of S&J providers by type of project and sub-study sample.** Across respondent-types, 2/3 or more of sub-study participants reported willingness to report physical or sexual victimization to an S&J provider, believed the case would be taken seriously, the survivor would be treated with respect, the survivor’s privacy would be protected, and the perpetrator would be punished by S&J actors. Broad patterns from the data indicate that 2/3 or more report expectations of survivor-centered practice across the S&J actors and that the women and female adolescents are more likely to report don’t know, indicating a lack of knowledge about the S&J actor. Some qualitative participants, mainly youth, specifically mentioned that they would go to police and other formal service providers over informal leaders and community members. “*In the past all the decision was made by the leader representative of the village and everyone used to trust their decision. But now there are Police and Mayor and Chief of ward. So people take these issues to them*” (Rupandehi, Female youth). The quantitative data support this as 89% of male youth and 88% of female youth report a willingness to report to the police. compared to 69% and 71% for a local leader, respectively. Most (87%) of the household survey sample and between 84% and 89% of the sub-study sample agreed or strongly agreed that police are trustworthy (**Table 4**).

**Table 4:** Perceptions of S&J providers, sub-sample.

|  | **Adult men**<br>**(N=221)** |  | **Male youth**<br>**(N=75)** |  | **Adult women**<br>**(N=243)** |  | **Female youth**<br>**(N=68)** |  |
| --- | --- | --- | --- | --- | --- | --- | --- | --- |
|  | % Yes | %<br>Don't<br>Know | % Yes | %<br>Don't<br>Know | %<br>Yes | %<br>Don't<br>Know | %<br>Yes | %<br>Don't<br>Know |
| Police are trustworthy | 89 | 4 | 89 | 3 | 84 | 5 | 88 | 1 |
| Citizens have role in law and order | 91 | 3 | 92 | 3 | 93 | 5 | 93 | 1 |
| **Police** |  |  |  |  |  |  |  |  |
| Willing to report | 89 | 0 | 89 | 0 | 83 | 1 | 88 | 0 |
| Case taken seriously | 79 | 2 | 81 | 7 | 78 | 5 | 84 | 6 |
| Survivor treated with respect | 80 | 2 | 84 | 7 | 76 | 6 | 88 | 6 |
| Privacy protected | 77 | 2 | 79 | 8 | 69 | 8 | 85 | 4 |
| Perpetrator Punished | 77 | 4 | 82 | 1 | 72 | 9 | 87 | 9 |
| Negative repercussions for reporting | 61 | 1 | 60 | 3 | 56 | 6 | 63 | 7 |
| **Judicial Committee** |  |  |  |  |  |  |  |  |
| Willing to report | 82 | 2 | 76 | 4 | 75 | 4 | 72 | 9 |
| Case taken seriously | 82 | 3 | 77 | 5 | 76 | 9 | 74 | 12 |
| Survivor treated with respect | 81 | 4 | 79 | 5 | 74 | 9 | 72 | 15 |
| Privacy protected | 77 | 4 | 75 | 5 | 69 | 11 | 63 | 16 |
| Perpetrator Punished | 77 | 5 | 72 | 6 | 67 | 13 | 69 | 16 |
| Negative repercussions for reporting | 59 | 3 | 63 | 3 | 57 | 7 | 54 | 16 |
| **Ward Chair** |  |  |  |  |  |  |  |  |
| Willing to report | 82 | 1 | 79 | 3 | 77 | 2 | 85 | 0 |
| Case taken seriously | 82 | 2 | 84 | 1 | 79 | 3 | 78 | 9 |
| Survivor treated with respect | 82 | 3 | 82 | 1 | 77 | 4 | 82 | 9 |
| Privacy protected | 77 | 4 | 81 | 3 | 71 | 7 | 71 | 9 |
| Perpetrator Punished | 77 | 5 | 81 | 1 | 68 | 8 | 77 | 9 |
| Negative repercussions for reporting | 62 | 3 | 59 | 1 | 57 | 4 | 60 | 9 |
| **Local Leader** |  |  |  |  |  |  |  |  |
| Willing to report | 77 | 2 | 69 | 7 | 70 | 6 | 71 | 6 |
| Case taken seriously | 77 | 4 | 71 | 8 | 72 | 9 | 69 | 12 |
| Survivor treated with respect | 76 | 5 | 75 | 8 | 69 | 9 | 71 | 12 |
| Privacy protected | 70 | 5 | 64 | 8 | 63 | 11 | 69 | 12 |
| Perpetrator Punished | 72 | 6 | 63 | 9 | 63 | 14 | 66 | 13 |
| Negative repercussions for reporting | 56 | 3 | 56 | 3 | 56 | 7 | 56 | 10 |
| **Women's Committee** |  |  |  |  |  |  |  |  |
| Willing to report | 73 | 2 | 75 | 3 | 76 | 3 | 75 | 3 |
| Case taken seriously | 77 | 5 | 76 | 1 | 79 | 5 | 78 | 7 |
| Survivor treated with respect | 75 | 5 | 80 | 1 | 79 | 6 | 77 | 7 |
| Privacy protected | 71 | 5 | 73 | 4 | 73 | 7 | 74 | 7 |
| Perpetrator Punished | 67 | 8 | 77 | 4 | 70 | 9 | 68 | 10 |
| Negative repercussions for reporting | 56 | 4 | 65 | 3 | 58 | 7 | 56 | 10 |

### Barriers to GBV help-seeking exist at multiple levels

While there is clear evidence that the police and other S&J providers are acceptable to approach, problems remain in availability, accessibility, speed and coordination, corruption, and perceived discrimination on the basis of socioeconomic status and caste. Further, the lack of female S&J providers and female representation, especially within community-focused remediation, remains a challenge to help-seeking. S&J providers identified the community’s lack of awareness as a major barrier to demand, which places survivors at risk of re-traumatization by seeking help from members of the community instead of the police, with out-of-school girls identified as a specifically vulnerable group. Because of this, many S&J providers prioritized community outreach and education as a means of increasing reporting. Stakeholders also suggested that more specifically targeted training of professionals and key community leaders would improve service provision and referral. Diverse stakeholders, including teachers, community members, and S&J providers, emphasized the importance of raising awareness of women’s and children’s rights and of resources available to provide support and assistance to victims of GBV.

> *“If a woman gets raped, she goes to 5 different places for help because of not knowing where to ask help from. Since her parents are uneducated, the child is also uneducated…she would first tell her parents then the parents would tell some of the influential persons of the village. They won’t directly come to the police station. That is the first time she gets raped. Then they would go to the ward chairperson which is the second time she gets raped. After that, they go to villagers and told them about being raped. It is like being raped 3 different times.” (Kapilvastu, Women’s cell)*

In some cases, community members were aware of S&J services for victims of GBV but were unable to fully access them due to distance and communication barriers. For example, a police officer from Rupandehi cited a lack of local language fluency among staff as a challenge to awareness-raising activities. Participants also stated that while there are police in every community, other organizations are prohibitively far from their communities. S&J providers cited distance and time spent travelling as barriers to victims who wish to escalate complaints within the judicial system.

Implementation of gender-sensitive practices is contingent upon having better-trained staff, sufficient female staff, and adequate budget available to provide services. S&J providers mentioned physical and human resources limitations as barriers to their capacity to address GBV in their communities, with the most commonly reported resource shortage being lack of staff. In terms of specific resources, a few S&J providers cited the need for safehouses to better protect victims and the need for private rooms for questioning at the police station. Additional budget to conduct trainings was also frequently mentioned.

Most negative opinions toward S&J providers from community members were related to bribery, politics, and discrimination, although it was often unclear whether such perceptions were based on lived experience. For example, one husband in Kapilvastu claimed, “If you have money, then only you will get help; otherwise, you will not.” However, help-seekers echoed these sentiments from their own experiences, with 3 out of 4 stating that they had faced discrimination based on gender and socioeconomic status.

> *“If the decision-maker is a male, all the judgments they make are focused on them and woman are deprived from proper judgment. Since we are female, they won’t listen to us. If a male walks into a decision-making panel, he won’t listen to the words of the women.” (Kapilvastu, Help-seeker)*

S&J providers also cited interference as common reasons for client complaints. In such cases, a perpetrator’s family may advocate on his behalf or involve influential others in the community to persuade the police to ignore the survivor’s complaint. “…*Sometimes, while resolving disputes and conflicts, we are pressurized by political parties or by other people too.”* (S&J provider, Siraha). S&J providers framed it as a hindrance to the execution of their duties that reduced public trust. Concomitantly, participants of all types across all districts cited it as a deterrence to reporting.

### Social and gender norms were perceived to be the largest barrier to help-seeking

These norms and associated cultural models fall into several categories (**Table 5**): 1) inequitable gender norms and related practices, including the practice of dowry, 2) the belief that marriage is sacred and life-long, 3) the belief that airing family problems bring shame and dishonor to the family, 4) the perception that while violence is undesirable, it is permissible when used to correct misbehavior, 5) the expectation that respect, obedience, and deference to elders’ decisions is paramount, and 6) the prioritization of family and social harmony which places the interests of the collective over those of individuals, particularly women.

**Table 5:**
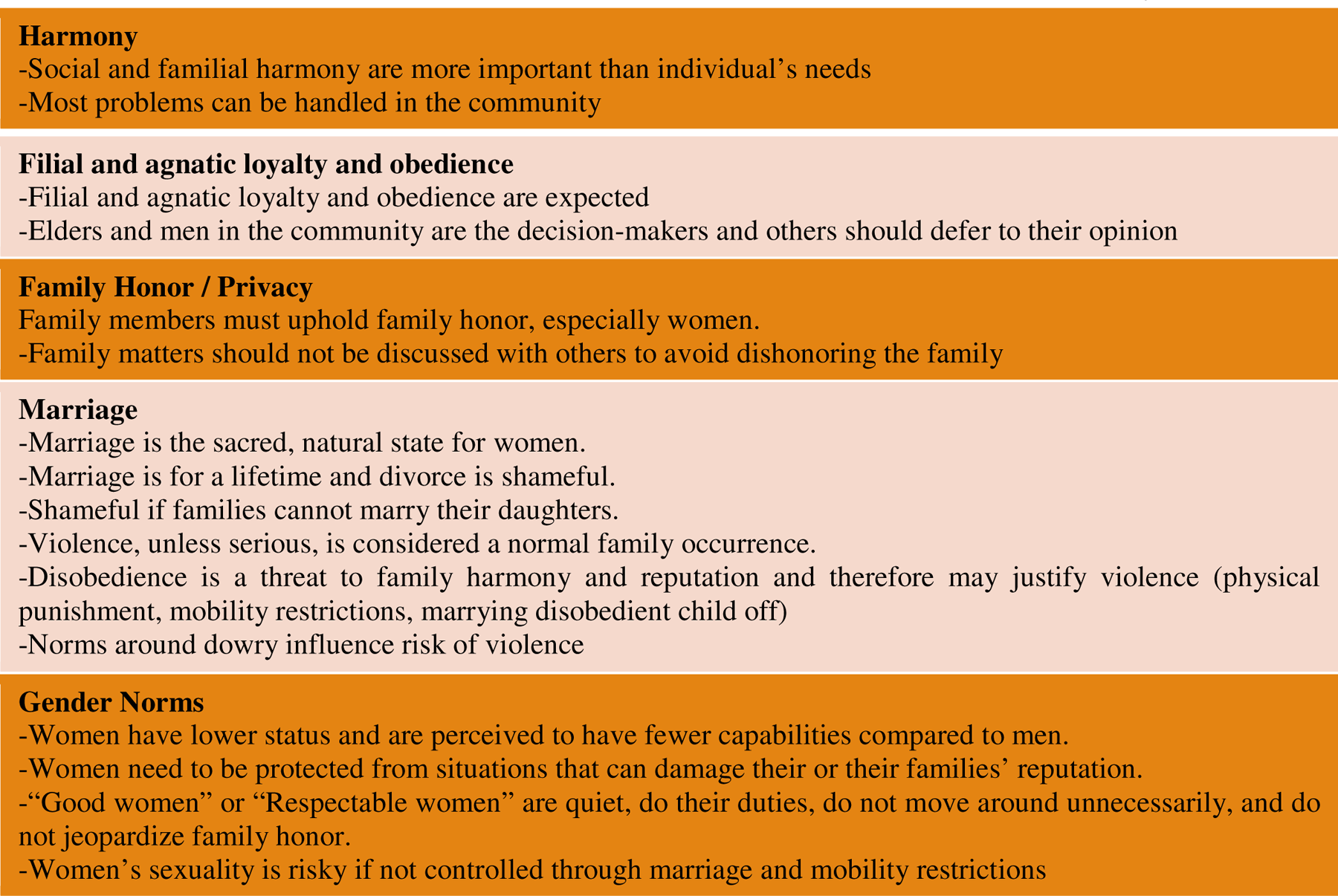
Social Norms, Gender Norms and Cultural Models Associated with Help-Seeking.

|  |
| --- |
| **Harmony**<br>-Social and familial harmony are more important than individual's needs<br>-Most problems can be handled in the community |
| **Filial and agnatic loyalty and obedience**<br>-Filial and agnatic loyalty and obedience are expected<br>-Elders and men in the community are the decision-makers and others should defer to their opinion |
| **Family Honor / Privacy**<br>Family members must uphold family honor, especially women.<br>-Family matters should not be discussed with others to avoid dishonoring the family |
| **Marriage**<br>-Marriage is the sacred, natural state for women.<br>-Marriage is for a lifetime and divorce is shameful.<br>-Shameful if families cannot marry their daughters.<br>-Violence, unless serious, is considered a normal family occurrence.<br>-Disobedience is a threat to family harmony and reputation and therefore may justify violence (physical punishment, mobility restrictions, marrying disobedient child off)<br>-Norms around dowry influence risk of violence |
| **Gender Norms**<br>-Women have lower status and are perceived to have fewer capabilities compared to men.<br>-Women need to be protected from situations that can damage their or their families' reputation.<br>-"Good women" or "Respectable women" are quiet, do their duties, do not move around unnecessarily, and do not jeopardize family honor.<br>-Women's sexuality is risky if not controlled through marriage and mobility restrictions |

A clear social order was described with household members, especially women deferring to men, and both men and women deferring to their elders, who are respected within the household and society. Despite younger generations being more educated than their parents and gaining more decision-making power, deference to men and elders remains the expectation. Girls are now more educated than in the past but are still raised to believe that the world is not a safe place for them and that they do not have the capability to speak or debate. This belief is also prevalent among men; as described by an S&J provider in Rupandehi, “almost 75% of males of this society think as such.” Women are socialized into a cultural model of inequality and are therefore considerably less powerful than men in the social hierarchy. Many women described limitations to their voice and mobility.

> *“It is a matter of fear to the women. Males are fearless but female has to fear. The males are able to understand and are aware and so they are fearless and are able to go out of their houses even at nighttime. But women are not able to do such things and it has never happened in our community as well.” (Saptari, Wife)*

It is considered uncommon for wives to challenge or raise their voice or hands against their husbands. Doing so is a violation of gender expectations and roles, widely perceived to be unacceptable and a trigger for violence. When responding to the vignette about a woman who raised her voice against her husband’s unreasonable behavior, most participants reported that “*she did not obey the rules imposed by society; she spoke loudly, she retorted. If one doesn’t act according to the rules of society, violence happens. This is what happens in our society in the Terai*.” *(Siraha, School management committee)*

Participants explained that because wives join their husbands’ households and become material and social assets for their marital families, they may lack support systems in their nuptial homes. While it is common that a woman returns to her natal family for support or refuge, the duration of refuge and the amount of support she receives is informed by how likely the woman and her family are to face social sanctions in the form of rumors and suspicions that the woman’s marriage is under threat. Divorce is financially untenable for most women, extremely shameful for her entire family, and perceived to be an unnatural state for women. Families have to be careful not to raise suspicions that a woman’s visit to her natal home has the potential to become permanent. These restrictions were reported to be especially strong in the Terai compared to hilly communities. “*If there’s some kind of argument with the husband then she will go to her mother’s house but eventually return to the husband’s house as it looks bad in the society*” *(Siraha, Male youth)*.

It is expected that most women will remain married, even after seeking help. Therefore, families face pressure to assist in a reconciliation or else risk having to provide lifelong refuge to their daughter and to live with the humiliation of this breakdown in expected social structure. Most participants reported that families will insist that their daughter not bring the shame and humiliation home with her. “*What will the community say about this and what will they think? Her parents aren’t well off and they won’t accept her; they will rather say “don’t come to us and don’t humiliate us, go back to your own house*” *(Siraha, S&J provider*). Given these challenges, especially the lack of familial support, participants widely perceived that most women would remain silent and bear the burden of violence.

While violence in general was very broadly defined and uniformly considered inappropriate across all sites, violence within the home was not considered a crime, especially when used to correct misbehavior. “*If [women and children] commit mistakes, punishing is not counted as an offence” (Rupandehi, Wife).* Husbands, heads of household, and teachers had the authority and responsibility to discipline errant behavior to maintain order. Our quantitative findings concur as sizable proportions of respondents believed that the use of physical violence by teachers against children, by parents against children, and by husbands against wives was acceptable within their community, especially in Madhesh Province (**Table 6**).

**Table 6:**
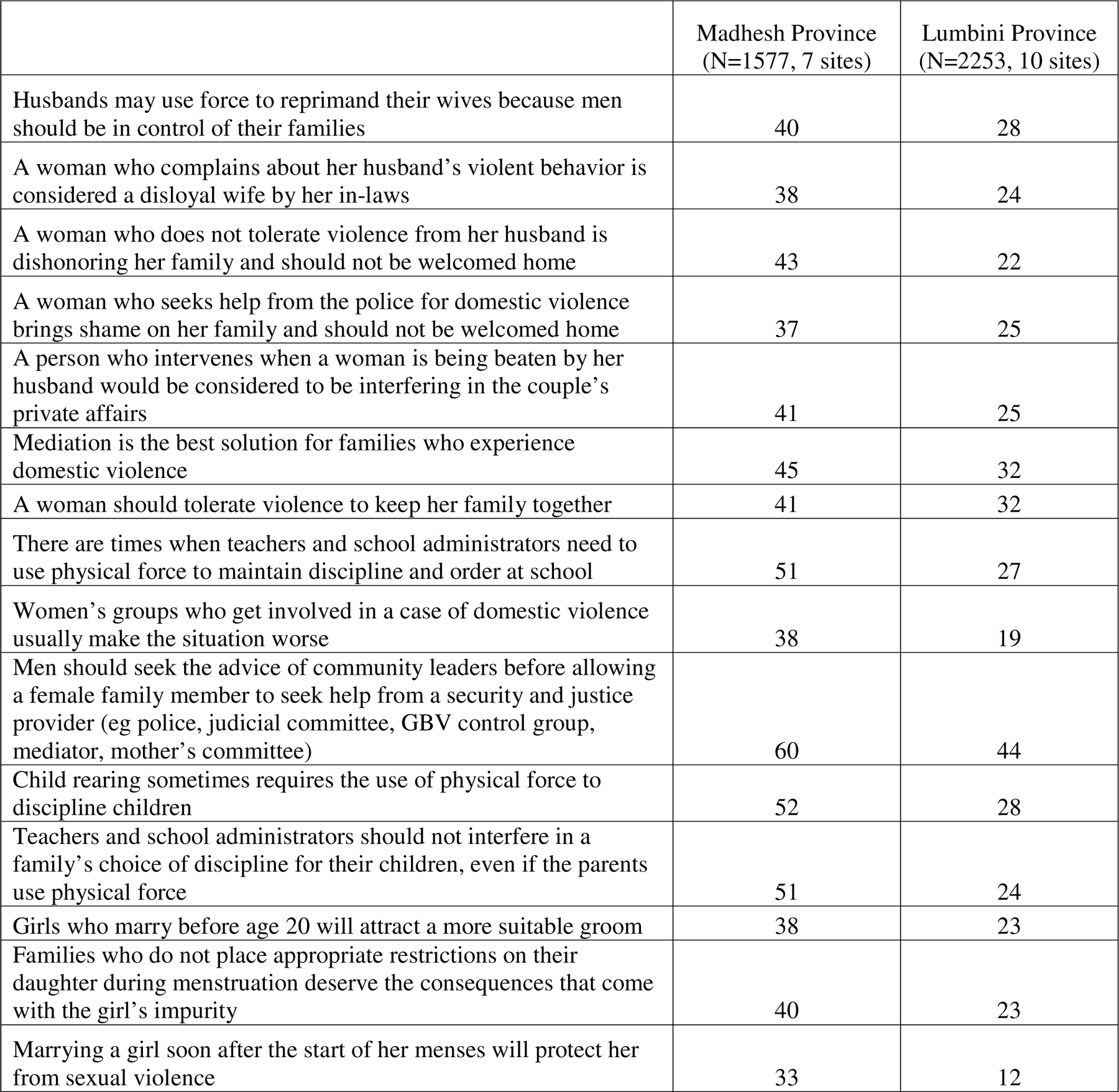
Mean percent reporting that a majority of their community holds these beliefs, household survey, by province.

|  | Madhesh Province<br>(N=1577, 7 sites) | Lumbini Province<br>(N=2253, 10 sites) |
| --- | --- | --- |
| Husbands may use force to reprimand their wives because men should be in control of their families | 40 | 28 |
| A woman who complains about her husband's violent behavior is considered a disloyal wife by her in-laws | 38 | 24 |
| A woman who does not tolerate violence from her husband is dishonoring her family and should not be welcomed home | 43 | 22 |
| A woman who seeks help from the police for domestic violence brings shame on her family and should not be welcomed home | 37 | 25 |
| A person who intervenes when a woman is being beaten by her husband would be considered to be interfering in the couple's private affairs | 41 | 25 |
| Mediation is the best solution for families who experience domestic violence | 45 | 32 |
| A woman should tolerate violence to keep her family together | 41 | 32 |
| There are times when teachers and school administrators need to use physical force to maintain discipline and order at school | 51 | 27 |
| Women's groups who get involved in a case of domestic violence usually make the situation worse | 38 | 19 |
| Men should seek the advice of community leaders before allowing a female family member to seek help from a security and justice provider (eg police, judicial committee, GBV control group, mediator, mother's committee) | 60 | 44 |
| Child rearing sometimes requires the use of physical force to discipline children | 52 | 28 |
| Teachers and school administrators should not interfere in a family's choice of discipline for their children, even if the parents use physical force | 51 | 24 |
| Girls who marry before age 20 will attract a more suitable groom | 38 | 23 |
| Families who do not place appropriate restrictions on their daughter during menstruation deserve the consequences that come with the girl's impurity | 40 | 23 |
| Marrying a girl soon after the start of her menses will protect her from sexual violence | 33 | 12 |

Qualitatively, respondents were consistent that problems within the family must be solved at home. Only after working without resolution with the family first and the larger community next should the justice sector be approached: that too with the support of the family and elders. Forty-four to 60% of survey respondents reported that a majority of their community believed that men should seek advice from community leaders before letting their female family member seek help from S&J providers (**Table 6**), which situates men as the gatekeepers for help-seeking. Women who reported violence to the police without first consulting family members and community leaders would risk losing community support and might incur blame for the violence according to the qualitative respondents according to the qualitative respondents. Across all respondent types, participants consistently believed that GBV could and should be dealt with within the community. Even most of the police indicated that after a report of domestic violence, they would work with the community to come to a solution. Therefore, it was nearly impossible to bypass the community even if one desired to do so. Our quantitative findings (**Table 6**) confirmed that most participants perceive a community norm that mediation is the best solution for GBV.

**The socially-approved approach to dealing with GBV, usually IPV, is some form of compromise and reconciliation even when seeking help from the police.** Reconciliation supports the notion of family and community harmony, often at the expense of the victim. While violence clearly detracts from family harmony, violating the social order risks even greater discord. Maintaining the established social order requires the cooperation of all families. Therefore, violating social norms could bring significant negative sanctions, ranging from backbiting to rumors and ostracizing. Across respondents (**Table 4**), most people are sensitive to these sanctions and are disinclined to violate them with over half to almost two-thirds believing that there would be negative repercussions for reporting across provider types.

> *“They are afraid of damaging their reputation in the neighborhood. They don’t want the news about the husband beating her to flash out in the community as they are newly married.” (Kapilvastu, Female youth)*

Parents and community leaders often enforce norms against reporting and encourage women and girls to tolerate violence to avoid shame and humiliation, which extends to the family and often to the entire village. Key reference groups, such as community members and leaders, often pressure women to reconcile with abusive husbands privately rather than involving police or other S&J providers.

> *“They used to say I am a bad woman. They taught my husband to kick me out of the house because I used to go to the police and the court to seek justice. They even used to say that I would never go to them for help if I was a good person.” (Rupandehi, Help-seeker)*

While the underlying norms were rather consistent across sites, there were also signs of differences in sensitivity to the sanctions. In all sites, while negative reactions to help-seeking for GBV from the community is the norm, there was discussion that some community members might react differently, especially if they are educated. Educated, “aware,” or “understanding” families were reported to be less sensitive to the sanctions associated with the norms and might have a more liberal approach to the problem. Across wards and participants, education was cited as a major factor increasing the likelihood of filing a formal report. Participants of all types stated that educated girls and women are more likely to report and that educated community members, including teachers and family members, would be more supportive of women and girls reporting GBV. This was explained by the greater knowledge and agency that educated individuals have, as well as weaker social norms against reporting among the educated.

> *“Those who are less aware think that she has damaged the reputation of not only the family but of the whole village by reporting it to the Police. Those who are aware think that it will bring positive change and will help protect other girls to fall as a victim of such incident.” (Saptari, S&J provider)*

Hilly communities were also recognized as having more liberal norms. As a father-in-law noted, changes are underway regarding women’s rights, which are perceived to be expanding in some settings.

## Discussion

### Descriptive norms

GBV was perceived to be widespread, especially child marriage, IPV, dowry-related domestic violence, and eve teasing. The higher percentage of persons perceiving forms of GBV to be more widespread in Madhesh province than in Lumbini province is in alignment with existing research on prevalence differences between these 2 provinces.(42, 43) The perception that GBV is widespread—especially when visible—contributes to its normalization as has been shown in prior research in Nepal.(44, 45) Enhancing the visibility of sanctions, including instances of community members publicly intervening, may help to shift expectations about its acceptability and normality.

### Disclosure

The relatively infrequent reports of IPV in the sub-study are somewhat consistent with, but lower than, IPV prevalence estimates generated within the broader study locality in prior IP-SSJ research.(46) The low prevalence of IPV in the sub-study suggests that it may be under-reported, possibly due to social desirability. However, comparable other data at this geographic level is lacking to be able to confirm if the reported estimate is an under-estimate. Data from other sites cannot be used to verify this study’s estimate given the great diversity of prevalence estimates of IPV demonstrated in research also at the tole-level in a different district.(47) The prevalence of child maltreatment in prior 12 months also does not have a direct comparison. It has been estimated that harsh physical punishment affects at least one in two children according to a nationally representative sample (48), suggesting that this study’s estimate that approximately 15% of the youth experienced child maltreatment (assessed as emotional, physical, and sexual violence, emotional and physical neglect, and witnessing IPV) is plausible.

Respondents were more likely to perceive child marriage as a crime compared to IPV. Research on child marriage in Nepal has shown the potential of legal change to bring social norms change if it is built on already occurring changes in behavior and attitudes and if there is sufficient awareness, along with competent, willing, and routine enforcement.(49) However, change in both forms of GBV has been modest. The median age at marriage has increased by only 1 year in the past decade(50) and physical and or sexual IPV in the prior 12 months has decreased 3% over a 5-year period,(50, 51) suggesting a lack of robust underlying behavior change on which the law might build. Further, while child marriage was more widely perceived to be a crime compared to IPV, it was described as a common practice. Laws against child marriage were reported to be inconsistent with social norms (particularly by S&J actors), and have been found to be circumvented(49) and unenforced,(52) along with laws against other forms of GBV, such as IPV(53) and *chhaupadi*.(54) An emphasis on social norms change as the prominent lever of behavior change seems warranted.

### Police and help-seeking from S&J providers

In alignment with prior research, respondents preferred community-based reconciliation to resolve issues of GBV (18, 21, 23) (22). The emphasis on reconciliation and prioritization of collective needs, which potentially protects social harmony and minimizes threats to family honor,(17, 18) can disadvantage female victims.(55) There is clear recognition across respondents that escalation of the issue to formal S&J providers is needed, especially when community-based solutions are ineffective, and the violence is repetitive and severe. S&J utilization in situations of severe violence is much more widely perceived to be acceptable in Nepal(56) and elsewhere.(57)

### Challenges to effective S&J service provision

There is some evidence that the police and other S&J providers are becoming more acceptable to approach and that community members perceive that victims will receive gender-sensitive and victim-centered services. Nevertheless, S&J service providers, GBV victims, and other respondents reported continued challenges with local S&J services. Similar findings from previous reports on IP-SSJ study areas suggest that perceptions of corruption and discrimination, interference, lack of female S&J officers, and accessibility are ongoing barriers to formal help-seeking (18, 21–23) that warrant continued redress if efforts to improve S&J acceptability and use for GBV are to be effective.

### Injunctive norms were perceived to be among the largest barriers to help-seeking

In a patriarchal, collectivist society,(58) there are strong expectations of agnatic and filial loyalty and deference. Extended-family living and enduring financial support and care of elders is predominant and has minimally changed over time in Nepal.(58) Evidence from this study suggests that there are some cracks in norms supporting elder decision-making and deference to community-mediation as the current generation of adults is more educated than the previous one. Nevertheless, the expectation remains that help-seeking should start at the family and the community and only when those interventions are ineffective should the issue be escalated to formal S&J services. In this study, it was widely expected that men should seek advice from family and community elders before assisting a female family member to seek justice. Prior research has examined this expectation, showing that while men seek counsel for conflict resolution, they may make their own decision, whereas women are much more tightly bound to the decisions made by men and community elders.(32) Men’s ability to make their own choices aligns with the qualitative findings of this study suggesting that while advice-seeking for conflict resolution is expected for men, they have the agency to decide their course of action whereas women do not.

It was clear from the study, that there were likely to be repercussions (negative sanctions) for formal help seeking. Help-seeking outside the family and community is perceived to bring dishonor on the husband and family, which, as shown in prior research in Nepal, is a violation of a woman’s familial duty.(22) Further, bypassing family and community mediation and reconciliation can result in strong social sanctions, increased blame placed on women, and a reduction in future social support. Highlighting successful survivor experiences,(32) addressing the S&J service challenges noted above, and working closely with community leaders and families to minimize the social consequences of help-seeking may collectively support a shift in expectations of social sanctions for help-seeking.

Respondents reported that violence was generally unacceptable and a threat to family harmony; however, respondents reported that violence used to correct disobedience, which threatens family order, harmony, and reputation was more acceptable, a reoccurring finding in prior research in Nepal(59–61) and globally (62). The salience of family honor throughout the study’s findings make it an important norm to link to violence prevention and the acceptability of help-seeking. Respondents perceived that while the perpetrator might be punished, or at least should be punished, women will still be blamed and dishonor will fall to her family.(22, 60) These linkages are not easy to disentangle and require community-based deliberation and stakeholder buy-in to shift the burden of dishonor from the victim to the perpetrator and to allow families to retain their honor by supporting female family members to live free of violence and seek formal justice when needed. While this outward-oriented behavior violates current practices of protecting family honor with secrecy, if reframed as a way to maintain family honor and harmony—values on which there is consensus—help-seeking and providing support for help-seeking might become more acceptable over time.

Girls and boys have access to educational opportunities that were not available to their parents and grandparents. Respondents believed that very early marriage is now generally considered to be harmful to girls and their well-being. However, boys and girls are still socialized into traditional gender roles,(63) with the expectation that men will protect and provide for their family and their parents, and women will take care of the home, her husband’s family, bear and raise children, and remain within the home unless necessary.(60) Greater support for the development of women’s and girls’ capabilities to counteract socialization into an inferior, dependent social status is needed to arm them with the skills they need to recognize and secure their rights(64) concomitant with strong male youth engagement to collectively transform gender norms.(22, 46, 60) In addition to families as the crucible of girls’ and boys’ socialization,(13) schools are an important venue for capability investment and norms change,(63) especially as teachers and other role models have been recognized as change agents for girls’ rights.(13) Engagement of female police officers and other female S&J actors in school, family, and community settings could provide additional, accessible role models for girls.

## Limitations

The study findings must be considered in light of its limitations. The chosen youth sub-study site was not suitable for a school-based assessment and last-minute changes in the field to restructure to a community-based sample were ineffective. The household survey was completed predominantly by married women, limiting assessment of norms’ perceptions to other socio-demographic groups. The sub-study represents only 1 site among 17.

## Conclusion

Normative change is clearly happening regarding child marriage. With greater education, there is slight evidence that deference to elders may be waning. Further, there is growth in awareness and perceived acceptability of formal S&J providers, although preferences for family- and community-based mediation remain strong. SAHAJ programming is already tapping into many of these opportunities and has been responsive to baseline findings in adjustments to their programming. Fully capitalizing on these opportunities and ensuring strong family-, school-, and community-based programming will bolster the project’s ability to detect normative and behavior change over its course, doubly so given its simultaneous focus on S&J social accountability and norms, which has been identified in prior IP-SSJ programming to be synergistic catalysts for change.(65)

## Supporting information

Online Supplement Table 1. Descriptive and injunctive norms, province mean, tole minimum and maximum.

## Data Availability

All data produced in the present work are contained in the manuscript

## Acknowledgements

We would like to offer special thanks to consortium leadership, management, and monitoring and evaluation technical experts at VSO Nepal (Jay Lal, Rachana Shrestha, Ratna Shrestha, Reena Chaudhary, Bikash Koirala) for expert guidance, project facilitation, and patience throughout the project, headquarter leadership and management at International Alert (Rabina Shrestha, Niresh Chapagain) for essential feedback throughout the project, Sudhindra Sharma and the team at Interdisciplinary Analysts for skillful and flexible data collection, staff at the Palladium Group (Bipa Shrestha, Danielle Stein, Coralie Blunier) for technical advice and tool sharing, and leadership (Karuna Onta, Mete Nielsen) and technical expertise (Moragh Loose) from the Foreign Commonwealth and Development Office in Nepal. We especially thank the study respondents for their time and responses without which the study would not be possible.

## Notes

### Competing Interest Statement

The authors have declared no competing interest.

### Funding Statement

Strengthening Access to Holistic Gender Responsive and Accountable Justice in Nepal was governed by a consortium led by Voluntary Service Overseas (VSO) with funding from the Foreign Commonwealth and Development Office when the funder was named the Department for International Development. The content is solely the responsibility of the authors and does not necessarily represent the official views of VSO or the funder.

### Author Declarations

Institutional Review Board at Emory University gave ethical approval for this work. Nepal Health Research Council gave ethical approval for this work.

