## Supplementary material for "Social Norms and Security and Justice Services for Gender-based Violence in Nepal: Programmatic Implications from a Baseline Mixed-Methods Assessment": Online Supplement Table 1. Descriptive and injunctive norms, province mean, tole minimum and maximum.

| **Online Supplement Table 1. Descriptive and injunctive norms, province mean, tole minimum and maximum.** | | | | | | |
| --- | --- | --- | --- | --- | --- | --- |
|  | Madhesh Province (N=1577, 7 sites) | | | Lumbini Province (N=2253, 10 sites) | | |
|  | mean | min | max | mean | min | max |
| **Descriptive norms (Percent reporting that the form of violence is widespread)** |  |  |  |  |  |  |
| Domestic violence | 57 | 33 | 69 | 34 | 9 | 50 |
| Dowry-related violence | 61 | 5 | 68 | 31 | 5 | 64 |
| Chaupadhi | 16 | 4 | 22 | 20 | 4 | 31 |
| Eve teasing | 49 | 40 | 56 | 49 | 42 | 58 |
| Sexual assault | 36 | 25 | 41 | 13 | 2 | 35 |
| Child marriage | 63 | 43 | 77 | 47 | 16 | 58 |
| Male-to-male violence | 45 | 40 | 54 | 24 | 10 | 42 |
| **Injunctive norms (Percent reporting that the majority of their community holds this belief)** |  |  |  |  |  |  |
| Husbands may use force to reprimand their wives because men should be in control of their families. | 40 | 24 | 61 | 28 | 0 | 60 |
| A woman who complains about her husband’s violent behavior is considered a disloyal wife by her in-laws. | 38 | 28 | 61 | 24 | 0 | 62 |
| A woman who does not tolerate violence from her husband is dishonoring her family and should not be welcomed home | 43 | 30 | 58 | 22 | 0 | 61 |
| A woman who seeks help from the police for domestic violence brings shame on her family and should not be welcomed home | 37 | 24 | 60 | 25 | 0 | 63 |
| A person who intervenes when a woman is being beaten by her husband would be considered to be interfering in the couple’s private affairs. | 41 | 25 | 58 | 25 | 2 | 61 |
| Mediation is the best solution for families who experience domestic violence | 45 | 25 | 62 | 32 | 24 | 58 |
| A woman should tolerate violence to keep her family together | 41 | 25 | 51 | 32 | 2 | 64 |
| There are times when teachers and school administrators need to use physical force to maintain discipline and order at school. | 51 | 36 | 62 | 27 | 5 | 61 |
| Women’s groups who get involved in a case of domestic violence usually make the situation worse. | 38 | 24 | 58 | 19 | 0 | 62 |
| Men should seek the advice of community leaders before allowing a female family member to seek help from a security and justice provider (e.g. police, judicial committee, GBV control group, mediator, mother’s committee) | 60 | 49 | 69 | 44 | 25 | 86 |
| Child rearing sometimes requires the use of physical force to discipline children. | 52 | 42 | 62 | 28 | 4 | 55 |
| Teachers and school administrators should not interfere in a family’s choice of discipline for their children, even if the parents use physical force. | 51 | 40 | 56 | 24 | 1 | 62 |
| Girls who marry before age 20 will attract a more suitable groom. | 38 | 22 | 46 | 23 | 0 | 59 |
| Families who do not place appropriate restrictions on their daughter during menstruation deserve the consequences that come with the girl’s impurity | 40 | 27 | 51 | 23 | 0 | 62 |
| Marrying a girl soon after the start of her menses will protect her from sexual violence | 33 | 24 | 42 | 12 | 0 | 40 |
